# Blink rates in patients with schizophrenia compared to healthy controls: A meta-analysis

**DOI:** 10.1101/2024.10.21.24315850

**Authors:** Johannes Reinau Windelborg Nielsen, Martin Dietz, Oskar Hougaard Jefsen

## Abstract

**Background:** Eyeblink rates vary with cognitive states and may be related to dopaminergic activity. Early reports have found elevated blink rates in patients with schizophrenia, possibly related to hyperdopaminergia, but findings appear inconsistent. We performed a systematic review and meta-analysis to evaluate the evidence regarding blink rates in patients with schizophrenia compared to healthy controls and the impact of medication.

**Methods:** We registered a protocol for the review on PROSPERO. We searched PubMed, SCOPUS, and Embase, performed title- and abstract-screening, full-text screening, data extraction, and risk-of-bias assessment. We calculated meta-level effect sizes, assessed effect size heterogeneity, and tested for small-study effects.

**Results:** We included 22 studies in the systematic review and of those, 19 in the meta-analysis. The studies included a total of 632 patients and 791 healthy controls. The risk of bias was high in all but two studies, particularly due to the lack of blinding and confounding by medication. Meta-analysis revealed an elevated blink rate in patients with schizophrenia (irrespective of medication status) compared to healthy controls (Hedges’ *g* = 0.48; 95% CI [0.13,0.82]). Stratified meta-analyses revealed elevated blink rates in unmedicated patients with schizophrenia compared with healthy controls (Hedges’ *g* = 0.83; 95% CI [0.34, 1.31]), but not in medicated patients compared with controls (Hedges’ *g* = -0.09; 95% CI [-0.64, 0.46]).

**Conclusion:** Blink rates are elevated in unmedicated, but not medicated, patients with schizophrenia compared with healthy controls. These findings motivate further study of the link between blink rates, central dopamine, and schizophrenia.

## 1. Introduction

The primary function of eyeblinks is to protect the ocular surface to maintain the precorneal tear film. Eyeblinks occur spontaneously or as a reflex induced by external stimuli, but can also be voluntary (Cruz, Garcia et al. 2011). Spontaneous eyeblinks typically occur 10-20 times per minute, but vary with age, cognitive processes, and activities (Jongkees and Colzato 2016). For example, higher blink rates are observed during fatigue (Caffier, Erdmann et al. 2003) and during conversation (Doughty 2001), while lower blink rates are observed during visually demanding activities such as reading (Bentivoglio, Bressman et al. 1997, Hoppe, Helfmann et al. 2018). Biologically, spontaneous eyeblink rates may be related to central dopaminergic activity (Jongkees and Colzato 2016); an idea sparked by observations in patients with altered dopaminergic function (Karson 1983). Early studies found elevated blink rates in patients with schizophrenia, associated with *hyper*dopaminergia (Karson 1983), and normalized blink rates during antipsychotic (i.e., antidopaminergic) treatment (Karson, Berman et al. 1983), and even abnormally low blink rates in patients experiencing antidopaminergic side effects (i.e., tardive dyskinesia or Parkinsonism) (Karson 1983). In contrast, studies have found abnormally low blink rates in patients with Parkinson’s disease, which is associated with *hypo*dopaminergia (Fitzpatrick, Hohl et al. 2012), an increase with dopaminergic treatment (Bologna, Fasano et al. 2012), and elevated blink rates in patients with hyperdopaminergic side effects (i.e., impulse control disorder, hallucinations) (Karson 1983). Moreover, experimental studies have demonstrated modulation of eyeblink rates by D1 and D2 receptor agonists and antagonists. However, the findings from human studies are mixed (Mohr, Sándor et al. 2005, Dang, Samanez-Larkin et al. 2017, Demiral, Manza et al. 2022).

Given the central role of the dopaminergic system in the treatment of schizophrenia, spontaneous eyeblink rates could potentially represent an important clinical marker and some studies have explored blink rates as a predictor of treatment response to antipsychotics (Karson, Bigelow et al. 1982, Lieberman, Kane et al. 1987, Chan, Hui et al. 2010). The strength of the evidence for elevated blink rates in patients with schizophrenia, however, remains uncertain. Some studies found no elevation in blink rates or even lowered blink rates in patients (Kitamura, Kahn et al. 1984, Kleinman, Karson et al. 1984) and findings may be affected by various sources of bias, such as blinding or confounding by medication). This calls for a systematic evaluation of the methodological quality of the literature, an exploration of the sources of heterogeneity, and a quantitative summary of the effect sizes.

To this end, we performed a systematic review and meta-analysis of studies investigating spontaneous eyeblink rates in patients with schizophrenia compared to healthy controls. We aimed to determine whether schizophrenia is associated with elevated blink rates and to assess the impact of medication and other sources of bias.

## 2. Methods

The protocol for this present study was preregistered on PROSPERO on July 8, 2023 (CRD42023440301).

### 2.1 Systematic literature search

We conducted systematic searches in PubMed, SCOPUS and EMBASE on June 25, 2023. The search string for PubMed was as follows: (“schizophrenia spectrum and other psychotic disorders”[MeSH Terms] OR “schizophreni*”[All Fields] OR “psychosis”[All Fields]) AND (“blink rate”[All Fields] OR “blinking”[MeSH Terms] OR “eyeblink*”[All Fields] OR “eye blink*”[All Fields]). Complementary search strings for EMBASE and SCOPUS searches are available in the supplementary material.

### 2.2 The screening process

Two investigators (JRWN and OHJ) performed the screening process in duplicate, independently of each other. Title- and Abstract screening was performed first, followed by Full Text Screening, using Covidence (www.covidence.org). At the end of both stages, disagreements on inclusion/exclusion were resolved by discussion.

### 2.3 Study selection

We included studies comparing patients with schizophrenia (according to DSM and ICD criteria) with healthy controls (no other psychiatric illnesses), restricting to publications in English. We included only studies reporting spontaneous eyeblinks rates. All comparative study designs were eligible. There were no restrictions regarding interventions, time frames, or year of publication.

### 2.4 Data extraction

Two investigators (JRWN and OHJ) performed the data extraction independently of each other using a duplicate, custom-made spreadsheet. The following study data were extracted:

I. Bibliometrics: Author and year of publication
II. Population: Sample size (N), female %, mean age, clinical characteristics (illness type, inpatient/outpatient), illness duration, medication status
III. Methodological characteristics: Method used (e.g. EOG, video observation), condition (e.g. Smooth Pursuit Eye Movement (SPEM), interview), lighting condition, time of day and year.
IV. Risk-of-bias: see below.
V. Quantitative results: spontaneous eyeblink rate means, standard deviations (or other error measures), or other sufficient statistics, from text or graphs using WebPlotDigitizer (Drevon, Fursa et al. 2017).

### 2.5 Risk-of-bias assessment

We employed a modified version of the Cochrane’s ROBINS-I-tool to assess the risk-of-bias of each study (Sterne, Hernán et al. 2016). We evaluated the following: (1) blinding of personnel to diagnostic status (2) Incomplete outcome data (3) Selective reporting (4) Bias due to confounding (e.g. by medication). Each part was evaluated in high/low/unsure. For item 4, medication-status was encoded as follows: medication-naïve = low, discontinued medication = unsure, medicated = high risk of bias. The overall risk-of-bias was defined as the highest risk-of-bias level on any of the 4 items. Disagreements were resolved through discussion. Risk of bias evaluation was visualized using robvis (McGuinness and Higgins 2021).

### 2.6 Meta-analysis

The meta-analysis was performed using STATA (StataCorp 2023). We calculated study-level effect sizes using Hedges’ g for the comparison of eyeblink rates between the patients with schizophrenia and healthy controls. If repeated measures were reported, we used data from the least confounded timepoint (i.e., medication-naïve < discontinued medication < medicated) for this comparison. Whenever possible, we compared eyeblink rates between unmedicated patients (i.e., discontinued medication or medication-naïve) versus controls and medicated patients versus controls, respectively, using similar methods. We evaluated effect size heterogeneity using Cochran’s Q-test and I^2^-statistics and calculated the summary effect sizes using DerSimonian-Laird random-effects meta-analysis (DerSimonian and Laird 1986). A Hedges’ g effect size > 0 indicates a higher mean blink rate in patients, while a Hedges’ g < 0 indicates a higher mean blink rate in the healthy controls. For studies reporting eyeblink rates from multiple conditions, we calculated a single effect size based on a pooled mean and standard deviation. For studies only reporting analytical (and no descriptive) statistics, these were converted to Hedges’ g using appropriate formulae (Lenhard & Lenhard, Psychometrica, 2016) (Supplementary Material). We investigated the impact of blinding on the effect size estimates using meta-level subgroup analysis.

## 3. Results

### 3.1 Included studies

Figure 1 displays a flowchart depicting the screening and inclusion of studies. After screening and assessment, 22 studies were included to the review. The main reason for exclusion during full-text screening was the lack of appropriate control group. Of the 22 studies included from full text screening, we excluded two studies from the meta-analysis due to insufficient statistical information (Cegalis and Sweeney 1979, Mackintosh, Kumar et al. 1983), and one study that measured maximum eyeblink rates rather than spontaneous eyeblink rates (Swarztrauber and Fujikawa 1998). The meta-analysis included 791 controls and 632 patients with schizophrenia.

**Figure 1.**
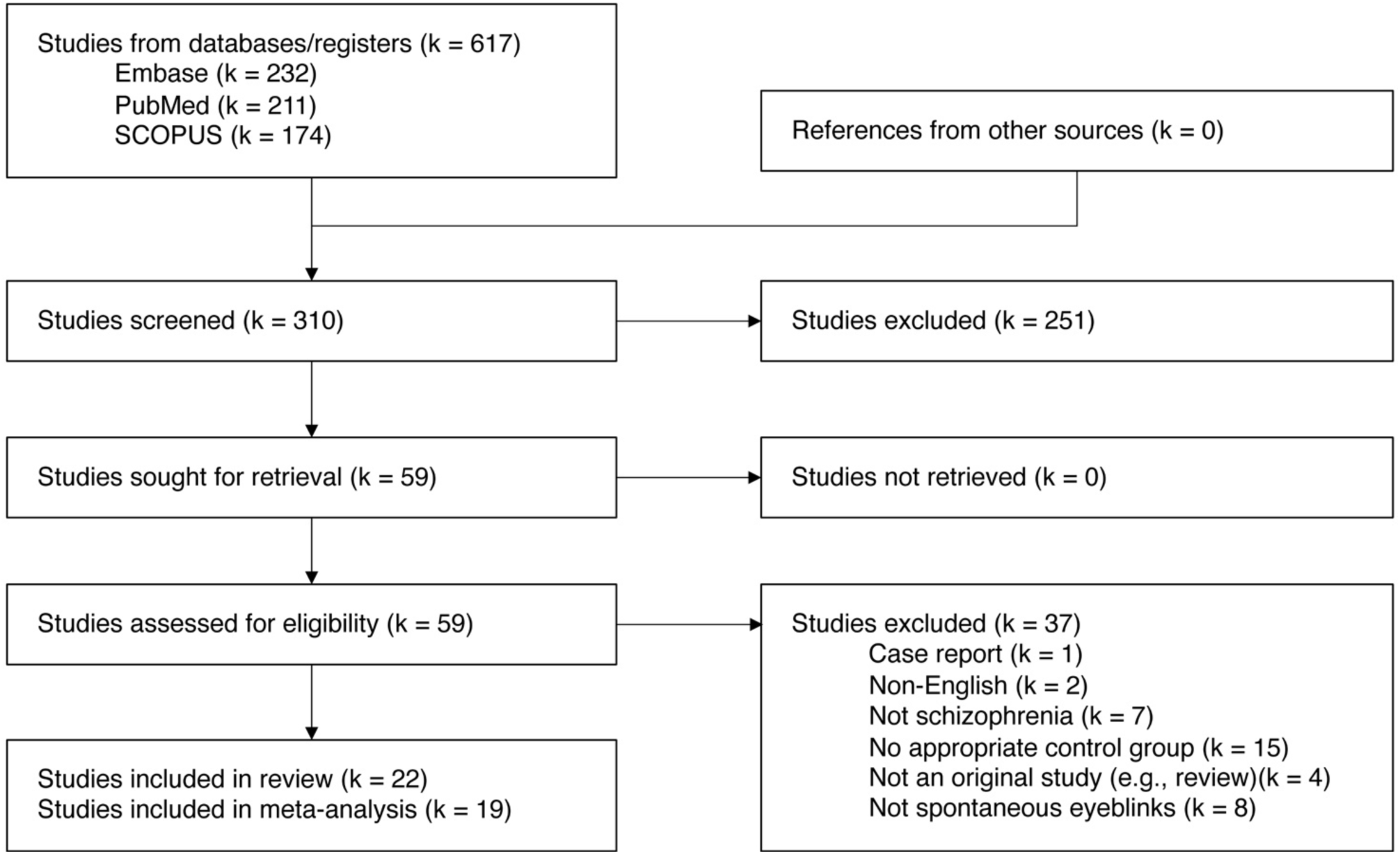
Flowchart describing the screening and inclusion of studies.

### 3.2 Participants

Table 1 describes the demographic and clinical characteristics, as well as medication status of the patients included in each study of our meta-analysis. Seven studies reported data from medication-naïve patients with four of these reporting separate data for the medication-naïve patients. Eleven studies reported data from patients with discontinued medication with nine of these studies reporting separate data for the patients with discontinued medication. Eighteen studies reported data from medicated patients with 14 of these studies reporting separate data for the medicated patients. The majority of studies reported data from inpatients with chronic schizophrenia.

**Table 1.** Participant characteristics for included studies.

|  | Healthy controls |  |  | Patients with schizophrenia |  |  |  |  |
| --- | --- | --- | --- | --- | --- | --- | --- | --- |
| Study | N | Sex,<br>% female | Age,<br>mean (SD) | N | Sex,<br>% female | Age,<br>mean (SD) | Medication status (n) | Clinical characteristics |
| Cegalis 1979 | 13 | 0% | 35.2 | 29 | 0% | Acute: 30.6<br>Chronic: 43.1 | Medicated and discontinued<sup>a</sup> | Acute (14) and chronic (15) |
| Freed 1980 | 19 | 26% | 24-56 | 30 | 30% | 21 to 46 | Medicated (19)<br>Discontinued (11)<sup>b</sup> | Inpatient |
| Karson 1981 | 39 | NR | 23-57 | 31 | 19% | 19-47 | Discontinued and medicated (same group of patients)<sup>b</sup> | Treatment-resistant inpatient |
| Karson 1983a | 54 | 37% | 36 (7) | 44 | 23% | 31 (7) | Discontinued and medicated (same group of patients)<sup>b</sup> | Treatment-resistant |
| Karson 1983b | 81 | NR | NR | 56 | NR | NR | Discontinued and medicated<sup>b</sup> | Chronic inpatient |
| Karson 1983c | 36 | 6% | 25 (6) | 27 | 22% | 31 (6) | Discontinued | Chronic inpatient |
| Mackintosh 1983 | 23 | NR | NR | 20 | NR | NR | Medicated | Chronic inpatient |
| Mueser 1984 | 21 | NR | NR | 11 | NR | NR | Medicated | Inpatient |
| Kitamura 1984 | 23 | 43% | 41.1 | 19 | 21% | 46.3 | Medicated | Chronic inpatient |
| Karson 1984 | 82 | 44% | 46 (10) | 55 | 24% | 31 (7) | Discontinued | Chronic inpatient |
| Kleinman 1984 | 81 | NR | NR | 55 | 36% | 19-51 | Discontinued and medicated<sup>a</sup> | Chronic inpatient |
| Helms 1985 | 12 | 42% | 29.8 (24-40) | 10 | 36%<sup>c</sup> | 30.1<sup>c</sup> | Medicated (7), discontinued (1) and medication-naïve (2)<sup>a</sup> | Inpatient |
| Mackert 1988 | 29 | NR | NR | 47 | NR | 31.8 | Medication-naïve (15) and discontinued (32)<sup>b</sup> | Inpatient |
| Mackert 1990 | 15 | 47% | NR | 15 | 47% | 30 (11) | Medication-naïve before (15) and after (8) medication<sup>b</sup> | Inpatient |
| Mackert 1991 | 28 | NR | NR | 26 | 50% | 30.7 (9.8) | Medication-naïve (8) and discontinued (18)<sup>b</sup> | Inpatient |
| Klein 1993 | 13 | 46% | 31 (6.5) | 13 | 31% | 33 (6.5) | Medicated | Remitted |
| Caplan 1994 | 53 | 17% | 9.3 (2.39) | 30 | 17% | 10.3 (1.53) | Medicated (16)<br>Discontinued (14)<sup>b</sup> | Inpatient (70%), outpatient (30%) |
| Adamson 1995 | 34 | 41% | 28 (3.6) | 40 | 40% | 29 (4.2) | Medication-naïve and medicated (same group of patients)<sup>b</sup> | Chronic (28)<br>Acute (12) |
| Jacobsen 1996 | 22 | 27% | 13.5 (2.2) | 17 | 41% | 14.5 (1.8) | Medicated (9);<br>Medication-naïve (8)<sup>a</sup> | Childhood-onset |
| Chen 1996 | 33 | 36% | 23.2 (4.2) | 40 | 48% | 23.7 (3.2) | Medicated | Inpatient |
| Swarztrauber 1998 | 35 | <10% | 40 | 23 | <10% | 42 | Medicated | Acute, subacute, chronic |
| Chan 2010 | 94 | 50% | 29 (9) | 93 | 55% | 31 (10) | Medication-naïve (48)<br>Medicated (45)<sup>a</sup> | Recruited at first psychotic episode |

<sup>a</sup> Results were not reported separately for each medication status.
<sup>b</sup> Results were reported separately for each medication status.
<sup>c</sup> Reported for a larger group of patients with psychosis, not for the patients with schizophrenia specifically.
NR = not reported.

### 3.3 Methodology

Supplementary Table S1. describes methodological characteristics of the included studies. The majority of studies measured eyeblink rates during interviews or visual tasks. Eyeblinks were typically detected by observation or EOG.

### 3.4 Risk of bias

The risk of bias assessment revealed a high risk of bias in all but two studies (Figure 2 and Supplementary Figure S1). The most common reason for high risk of bias was confounding by medication. Moreover, investigators were not blinded in more than 60% of studies. Most studies supplied complete outcome data and were not considered to show selective reporting.

**Figure 2.**
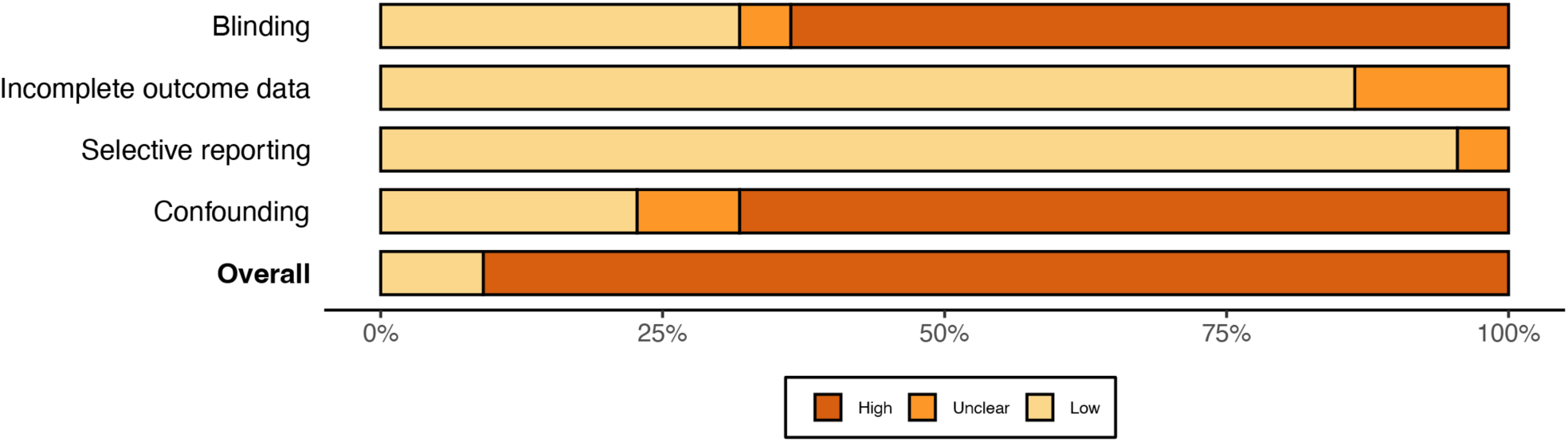
Summary of risk-of-bias-assessment of the included studies (k = 22)

### 3.5 Meta-analysis – patients versus healthy controls

Patients with schizophrenia showed significantly higher blink rates compared to healthy controls (Hedges’ g = 0.48; 95% CI [0.13,0.82]) as shown in Figure 3. There was a high degree of effect size heterogeneity (I^2^ = 88.40%). We found no evidence of small-study effects using Egger regression (*p* = 0.35) (Supplementary Figure S1). Finally, we found no statistically significant difference between the summary effect sizes from studies with vs. without blinded outcome assessments (χ^2^ = 0.19, *p* = 0.666).

**Figure 3.**
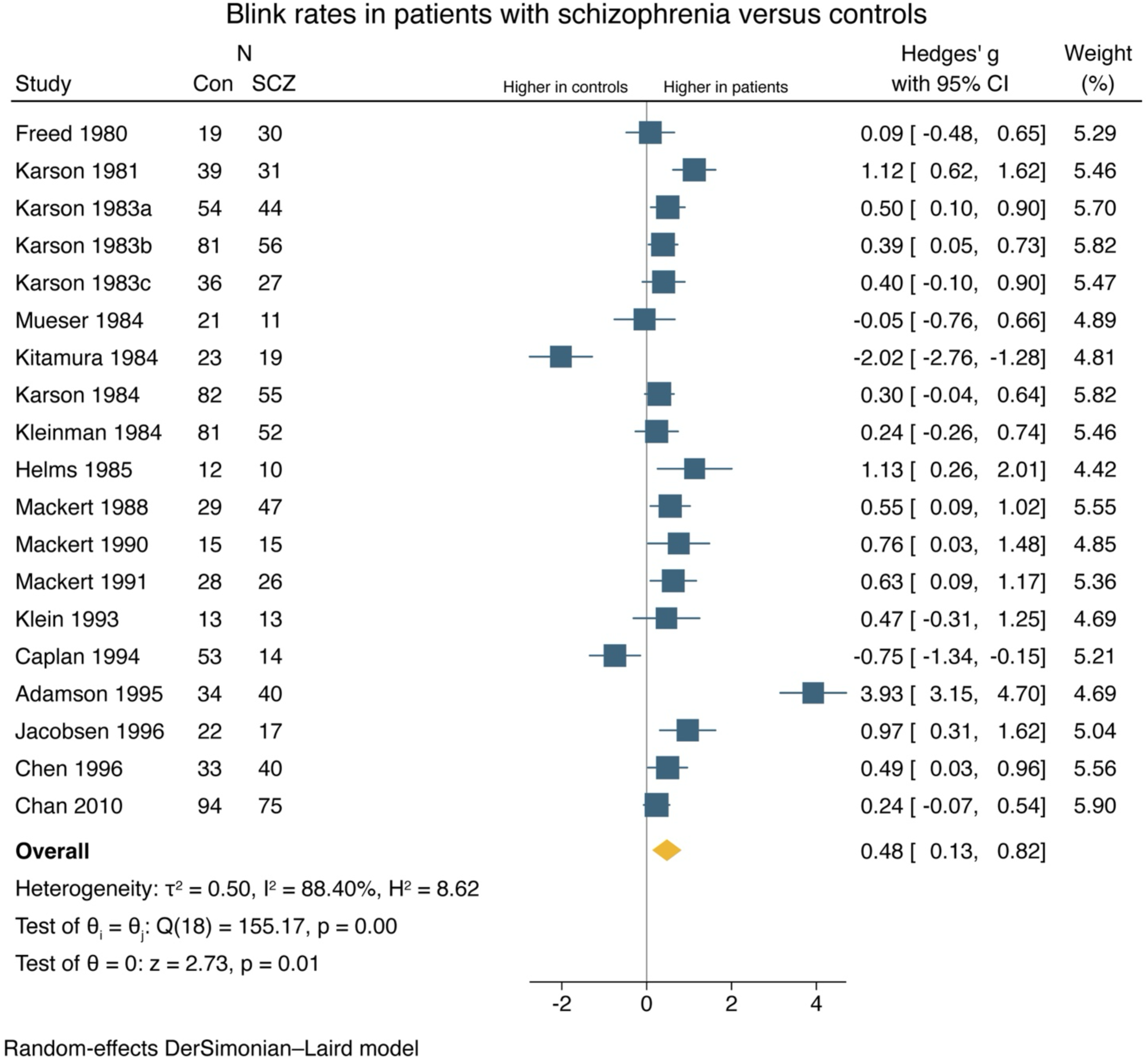
Forest plot of effect sizes (patients with schizophrenia versus healthy controls) (k = 19).

### 3.6 Meta-analysis – medication status

Unmedicated patients (including both medication-naïve patients and patients discontinued from medication) showed significantly higher blink rates compared to healthy controls (Hedges’ g = 0.83; 95% CI [0.34,1.31]) (Figure 4), albeit with large effect size heterogeneity (I^2^ = 90.57%). Here, we found evidence of small-study effects using Egger regression (*p* = 0.002) (Supplementary Figure S3). In contrast, patients on antipsychotic medication showed no increase/decrease in blink rates compared to healthy controls (Hedges’ g = -0.09, 95% CI [-0.64, 0.46]) (Figure 4). In this analysis, we also found a high degree of effect size heterogeneity (I^2^ = 91.22%), but no evidence of small-study effects (*p* = 0.64) (Supplementary Figure S4). Finally, we conducted a small meta-analysis of three studies comparing only medication-naïve patients with healthy controls. We found no increase or decrease in blink rates compared with controls (Hedges’ g = 1.74, 95% CI [-0.30, 3.78]) (Supplementary Figure S5).

**Figure 4.**
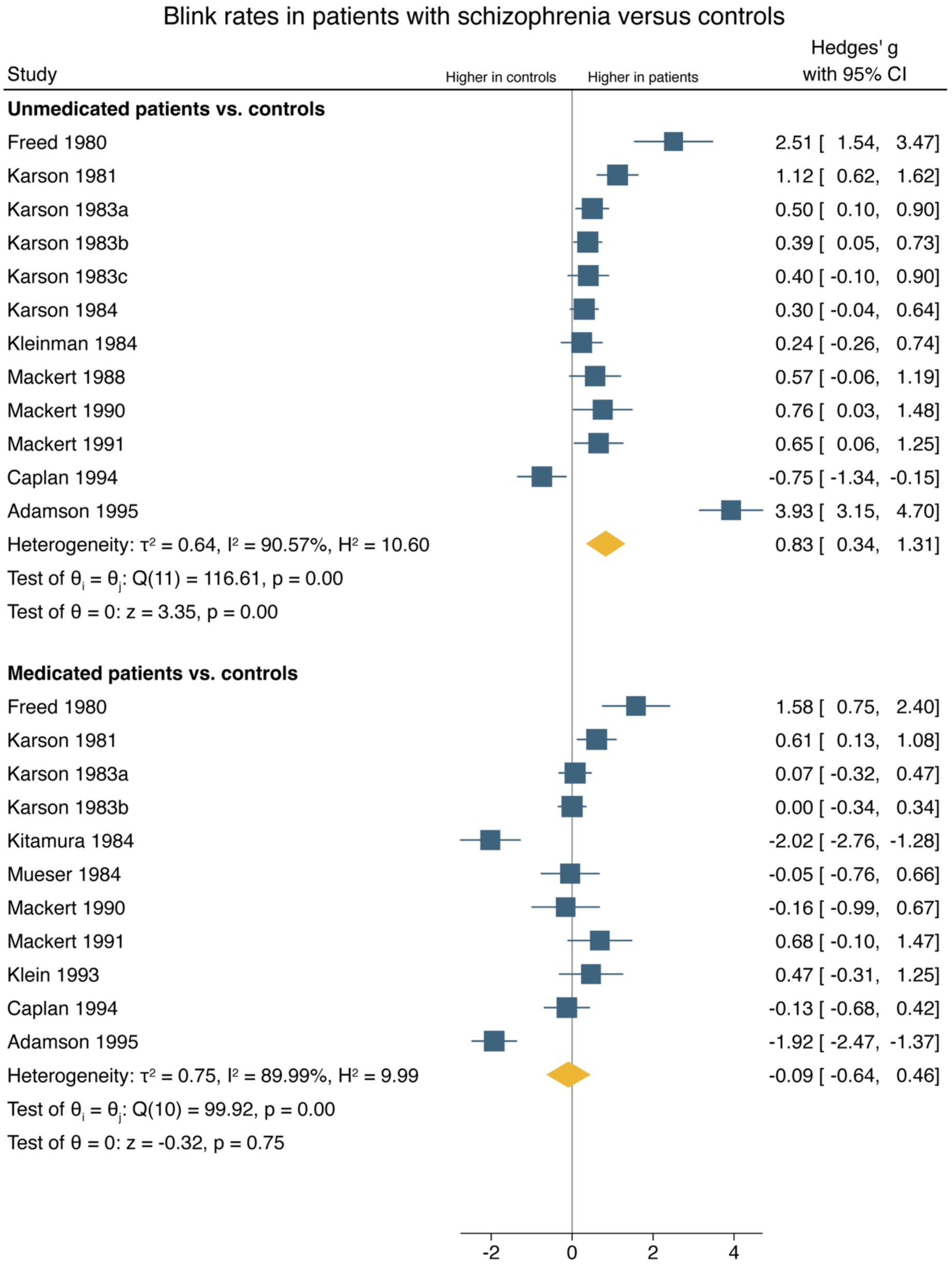
Forest plot of effect sizes (patients with schizophrenia versus healthy controls) stratified for medication status.

## 4. Discussion

We conducted a systematic review and meta-analysis of spontaneous eyeblink rates in patients with schizophrenia compared to healthy controls. Overall, patients with schizophrenia displayed elevated eyeblink rates compared to healthy controls. The elevation was pronounced in unmedicated patients but absent in medicated patients. The risk of bias was high in all but two studies, mainly due to lack of blinding and/or confounding by medication.

A previous narrative review by Jongkees and Colzato (Jongkees and Colzato 2016) provided a qualitative overview of eyeblink rates in clinical populations, including patients with schizophrenia, and noted consistencies and discrepancies between studies. Our study adds a systematic risk of bias assessments and provides meta-analytical evidence for the elevation of blink rates in schizophrenia. Elevated blink rates have been suggested to represent a proxy of striatal hyperdopaminergia in schizophrenia – a main treatment target for antipsychotic medication – and could thus hold clinical potential. Blink rates may thus potentially be considered an easily obtainable, but imprecise, marker of central dopaminergic function. However, it should be noted that the link between blink rates and central dopaminergic activity remains disputed (Dang, Samanez-Larkin et al. 2017). Future studies may determine the mechanisms (dopaminergic or other) underlying blink rate elevation in schizophrenia and study its clinical utility

Certain methodological limitations guide the interpretation of our findings, some of which pertain to the included studies, and some pertaining to our study. As revealed in the risk of bias assessment, the majority of the included studies had a high risk of bias, particularly due to lack of blinding and confounding by medication. The lack of blinding in personnel could potentially lead to an over- or underestimation of the number of eyeblinks, when the recording method was not automatic. As only three studies reported data from medication-naïve patients, we are unable to fully exclude confounding by medication in the present results. On a meta-analytic level, we found a high degree of effect size heterogeneity, even after controlling for medication status, indicating the presence of unidentified sources of between-study variance. Another limitation pertains to the risk of publication bias. We did not detect any small-study effects; however, we are unable to completely disregard publication bias.

In conclusion, schizophrenia is associated with higher spontaneous blink rates in unmedicated patients, but not in medicated patients. Further studies on the link between blink rates and dopamine, and between blink rates and treatment response, are needed to determine the clinical utility of blink rate assessments in psychiatry. We suggest that future studies apply blinded assessment, focus on medication-free patients, and use automated detection of eyeblinks to avoid bias.

## Data Availability

All data produced in the present study are available upon reasonable request to the authors

