## Supplementary material for "Blink rates in patients with schizophrenia compared to healthy controls: A meta-analysis"

**Affiliations**
[1] Center of Functionally Integrative Neuroscience, Department of Clinical Medicine, Aarhus University, Aarhus, Denmark.
[2] Psychosis Research Unit, Aarhus University Hospital – Psychiatry, Aarhus, Denmark.

**Corresponding author**
Oskar Hougaard Jefsen


### Supplementary Methods

**Search strings**

PubMed:

("schizophrenia spectrum and other psychotic disorders"[MeSH Terms] OR "schizophreni*"[All Fields] OR "psychosis"[All Fields]) AND ("blink rate*"[All Fields] OR "blinking"[MeSH Terms] OR "eyeblink*"[All Fields] OR "eye blink*"[All Fields])

EMBASE:

('schizophreni*' OR 'psychosis'/exp) AND ('eye blink*' OR 'eyeblink*' OR 'blink rate*')

SCOPUS:

TITLE-ABS-KEY ( "schizophreni*" OR "psychosis" ) AND TITLE-ABS-KEY ( "eye blink*" OR "eyeblink*" OR "blink rate*" )

**Conversion of effect sizes: Kleinman 1984**

Reported in the paper:

> Unmedicated patients with normal ventricular brain ratio (NVBR) (n = 42) vs. controls (n = 81). Results of Mann-Whitney test; U = 1388.5.

Effect size calculator on https://www.psychometrica.de/effect_size.html#transform yields Cohen's d-value of 0.304, which is further converted below:

$$g = d * \left(1 - \frac{3}{4 * (n_1 + n_2 - 2) - 1}\right)$$

$$g_{NVBR} = 0.304 * \left(1 - \frac{3}{4 * (81 + 42 - 2) - 1}\right) = 0.302$$

$$SE_g = \sqrt{\frac{n_1 + n_2}{n_1 n_2} + \frac{g^2}{2(n_1 + n_2)}}$$

$$SE_{g_{NVBR}} = \sqrt{\frac{81 + 42}{81 * 42} + \frac{0.304^2}{2(81 + 42)}} = 0.191$$

Reported in the paper:

> Unmedicated patients with larger ventricular brain ratio (LVBR) (n = 11) vs. controls (n = 81). Results of Mann-Whitney test; U = 394.

Effect size calculator on https://www.psychometrica.de/effect_size.html#transform yields Cohen's d-value of 0.129, which is further converted below:

$$g = d * \left(1 - \frac{3}{4 * (n_1 + n_2 - 2) - 1}\right)$$

$$g_{LVBR} = 0.129 * \left(1 - \frac{3}{4 * (81 + 11 - 2) - 1}\right) = 0.128$$

$$SE_g = \sqrt{\frac{n_1 + n_2}{n_1 n_2} + \frac{g^2}{2(n_1 + n_2)}}$$

$$SE_{g_{LVBR}} = \sqrt{\frac{81 + 11}{81 * 11} + \frac{0.128^2}{2(81 + 11)}} = 0.321$$

We then calculate a weighted average of the two effect sizes (weighted by the SE)

$$g_{total} = \frac{g_{NVBR} * SE_{g_{NVBR}}^{-1} + g_{LVBR} * SE_{g_{LVBR}}^{-1}}{SE_{g_{NVBR}}^{-1} + SE_{g_{LVBR}}^{-1}}$$

$$g_{total} = \frac{0.302 * 0.191^{-1} + 0.128 * 0.321^{-1}}{0.191^{-1} + 0.321^{-1}} = 0.237$$

And calculate the overall SE as a simple average of the two subgroup SEs.

$$SE_g = \frac{SE_{g_{NVBR}} + SE_{g_{LVBR}}}{2}$$

$$SE_g = \frac{0.191 + 0.321}{2} = 0.256$$

**Conversion of effect sizes: Klein 1993**

Reported in the paper: Patients (n = 13) and controls (n = 13). Result of ANOVA: F = 1.4.

Conversion into Hedges' g below:

$$d = \sqrt{F\left(\frac{n_1 + n_2}{n_1 n_2}\right)\left(\frac{n_1 + n_2}{n_1 + n_2 - 2}\right)}$$

$$d = \sqrt{1.4\left(\frac{13 + 13}{13 * 13}\right)\left(\frac{13 + 13}{13 + 13 - 2}\right)} = 0.483$$

$$g = d * \left(1 - \frac{3}{4 * (n_1 + n_2 - 2) - 1}\right)$$

$$g = 0.483 * \left(1 - \frac{3}{4 * (13 + 13 - 2) - 1}\right) = 0.468$$

$$SE_g = \sqrt{\frac{n_1 + n_2}{n_1 n_2} + \frac{g^2}{2(n_1 + n_2)}}$$

$$SE_g = \sqrt{\frac{13 + 13}{13 * 13} + \frac{0.468^2}{2(13 + 13)}} = 0.398$$

#### Supplementary Tables

| Supplementary Table S1. Measurement of spontaneous eyeblinks | | |
|---|---|---|
| **Study** | **Condition** | **Detection method** |
| Cegalis 1979 | SPEM (four conditions) | Eye tracking |
| Freed 1980 | Interview | Observation |
| Karson 1981 | Interview | Observation |
| Karson 1983a | Interview | Observation |
| Karson 1983b | Speech | NR |
| Karson 1983c | Interview | Observation |
| Mackintosh 1983 | Interview | Video recording |
| Mueser 1984 | Interview | Observation |
| Kitamura 1984 | Interview | Video recording |
| Karson 1984 | Interview | Observation |
| Kleinman 1984 | Interview | Observation |
| Helms 1985 | Visual task | Observation |
| Mackert 1988 | Visual task | EOG |
| Mackert 1990 | Visual task | EOG |
| Mackert 1991 | Visual task | EOG |
| Klein 1993 | Auditory task and SPEM | EOG |
| Caplan 1994 | Listening, conversation, and verbal recall | Video recording |
| Adamson 1995 | Interview | Observation |
| Jacobsen 1996 | SPEM | Observation |
| Chen 1996 | Listening to music; auditory task | Observation |
| Swarztrauber 1998 | Rest; cognitive testing, phototic stimulation | EEG |
| Chan 2010 | Listening to music | Observation |

EEG = electroencephalography. EOG = electrooculography. NR = not reported. SPEM = Smooth pursuit eye movement task.

#### Supplementary Figures

**Supplementary Figure S1. Traffic light plot of risk of bias evaluations.**

**Supplementary Figure S2. Funnel plot (blink rates in patients with schizophrenia versus controls).**

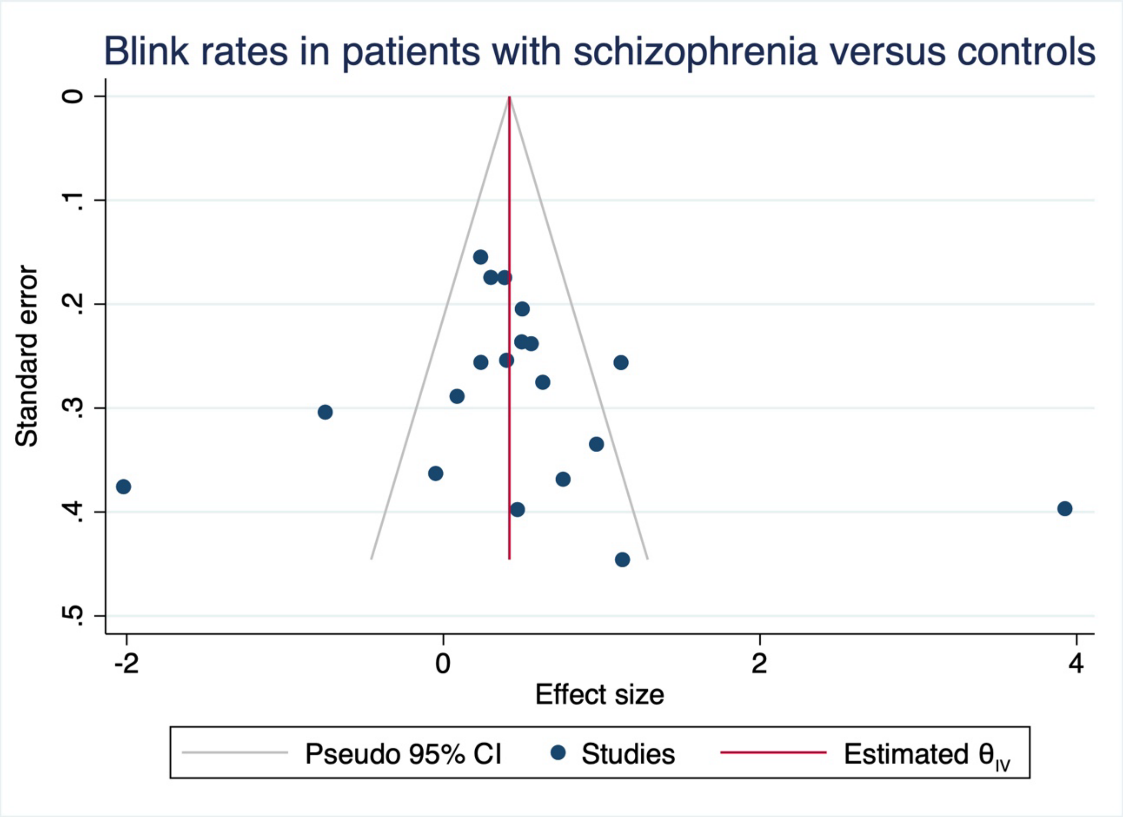

**Supplementary Figure S3. Funnel plot (unmedicated patients versus controls).**

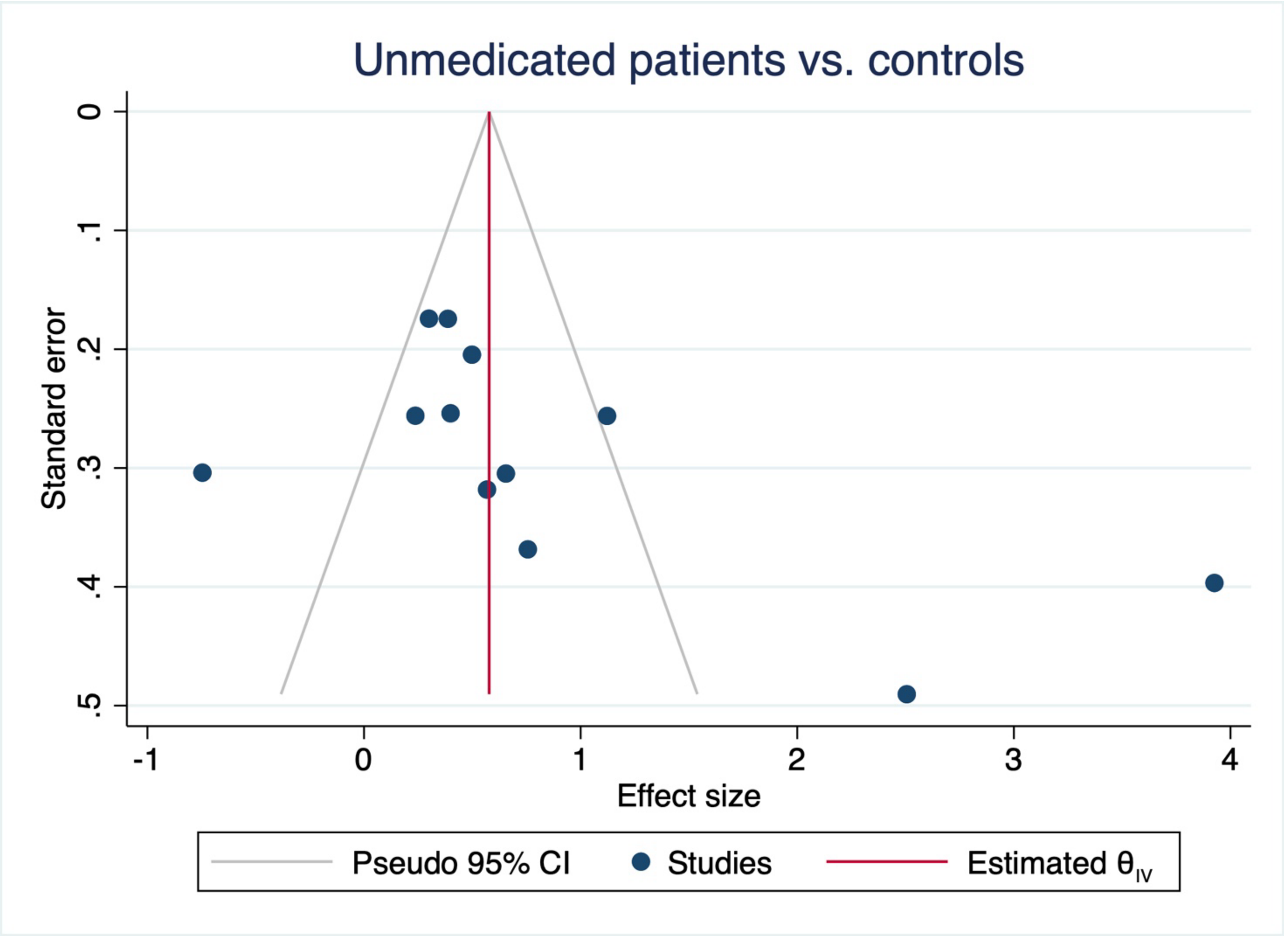

**Supplementary Figure S4. Funnel plot (medicated patients versus controls).**

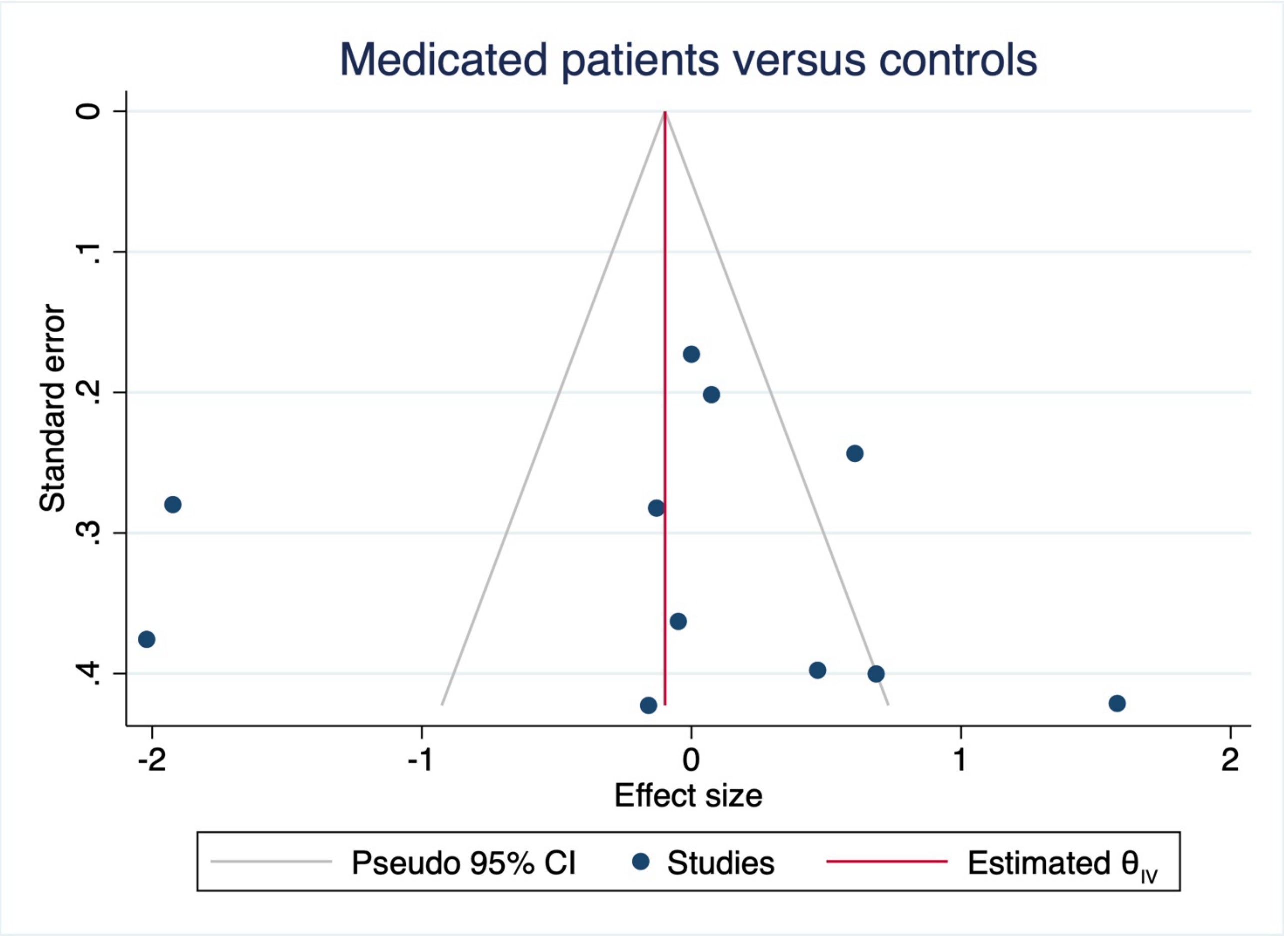

**Supplementary Figure S5. Forest plot (medication-naïve patients versus controls).**

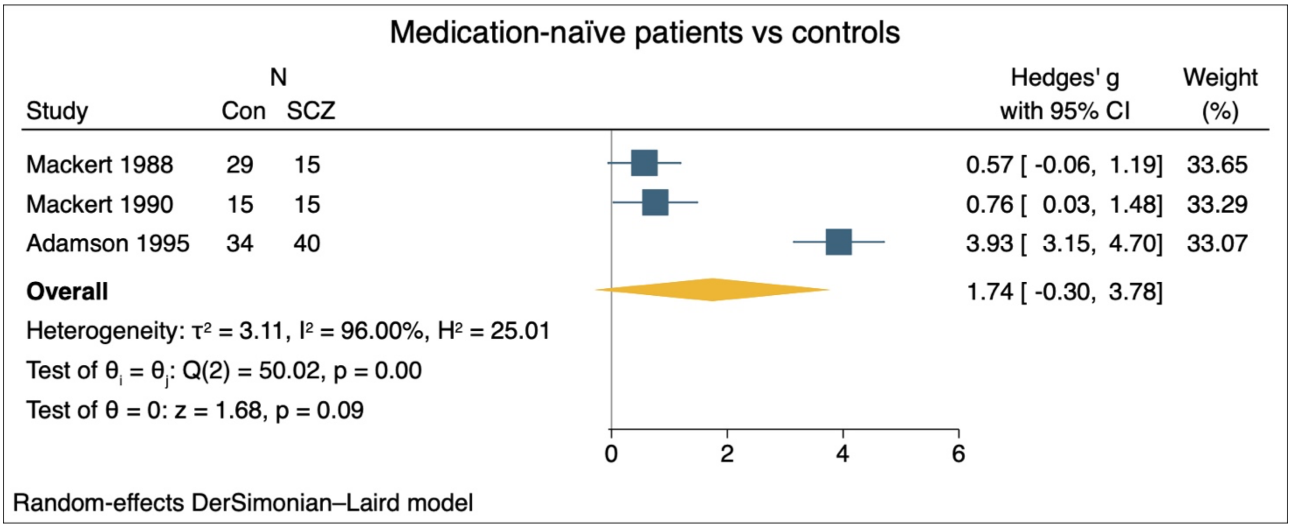
